# Exploring problematic internet use and gaming in young adults with autism spectrum disorder

**DOI:** 10.1101/2022.09.16.22279979

**Authors:** Claude L. Normand, Marisa H. Fisher, Stéphanie-M. Fecteau, Kelly Tremblay, Evelyne Roy, Marie-Hélène Poulin

**Affiliations:** Département de psychoéducation et de psychologie, Université du Québec en Outaouais, Gatineau, QC, Canada; Department of Counseling, Educational Psychology and Special Education, Michigan State University East Lansing, MI; Département de psychoéducation et de psychologie, Université du Québec en Outaouais, Saint-Jérôme, QC, Canada; Département des sciences du développement humain et social, Université du Québec en Abitibi Témiscamingue, Rouyn-Noranda, QC, Canada

**Author notes:** **Corresponding author contact information** Claude L. Normand, Département de psychoéducation et de psychologie, Université du Québec en Outaouais, Case postale 1250, Succursale Hull, Gatineau, Qc Canada J8X 3×7; Poste: 2242; Courriel.

**Keywords:** autism spectrum disorder, gaming disorder, internet use, video games, addiction, young adult

## Abstract

Characteristics of autism spectrum disorder (ASD) could lead individuals with ASD to spend large amounts of time on internet and potentially becoming addicted. Sixty-five individuals with ASD (mean age = 23.77, SD = 4.3) completed an online survey measuring compulsive internet use and gaming disorder. Six participants (9.3%) had problematic internet use, while only two (3%) scored above cut-off for gaming disorder. Exploratory analyses found no differences according to gender, age, employment or co-occurring diagnoses of anxiety, ADHD or depression. Our data suggest that problematic internet could be more frequent than gaming disorder in Canadian young adults with autism and that these issues should be investigated further.

## Introduction

The proliferation of mobile information and communication technologies (ICT), the widespread availability of Wi-Fi in public settings, as well as the social contact restrictions during the Covid-19 pandemic have created a context where people spend more time than ever online. This combination of factors has triggered concerns regarding the possibility of an increase in problematic internet use, among young people especially. Although “normal” or adequate internet use is difficult to define, since it evolves with changes in the technologies and their applications, it is important to note that problematic internet use or addiction cannot be assessed only by the number of hours spent online daily (Griffiths, 2018; Király et al., 2017).

### Defining the Concepts of Problematic Internet Use and Gaming

The concept of internet addiction (IA) was made popular by Kimberly Young in 1998, when she published the *Internet Addiction Test*, a measure that assesses internet usage in terms of the diagnostic symptoms of substance use disorder, including degree of preoccupation, inability to control use, extent of hiding or lying about internet use, and continued online use despite negative consequences of behavior (Young, 1998). Since then, many clinicians and researchers have attempted to define and assess this non-substance related disorder (Bergmark et al., 2016; Cash et al., 2012; Czincz & Hechanova, 2009; Griffiths, 2018; Hou et al., 2019; Kardefelt-Winther et al., 2017; Kuss & Griffiths, 2012; Ryding & Kaye, 2017; Sussman et al., 2018). Overall, they agree that the *impact* of the amount of time spent online (e.g. losing sleep, getting into arguments, neglecting one’s obligations) and other signs of addiction (e.g. preoccupation and withdrawal symptoms) are deemed more important to distinguish problematic internet use (or addiction) from “normal” or excessive use.

Problematic internet use can also be circumscribed to online video games. Like internet addiction, defining gaming disorder (GD) has been a source of debate among researchers (Griffith & Meredith, 2009; Király et al., 2014; Lehenbauer-Baum & Fohringer, 2015; Montag et al., 2019b; Sussman et al., 2018). Although the *DSM-5* (APA, 2013) has classified internet gaming disorder (IGD) as an “emerging” condition, the World Health Organisation (WHO) formally recognized gaming disorder (GD) in its 11^th^ edition of the

*International Classification of Diseases* (ICD-11; WHO, 2019). Three main criteria (present for at least 12 months) characterize GD: impaired control over gaming, continuing or increasing time spent gaming despite negative consequences, and neglecting other hobbies in favour of gaming. Meanwhile, the DSM-5 defines IGD by nine criteria, closely resembling those for substance use disorder: preoccupation with games, withdrawal symptoms, tolerance, a lack of self-control concerning video game play, a loss of interest in hobbies, continuation and escalation despite consequences, deception around the amount of time spent gaming, use of gaming to escape negative feelings and negative consequences including risking or losing a job or relationship due to gaming (Engelhardt et al., 2017; Petry et al., 2014).

It has been suggested that excessive gaming may be transitory and not necessarily lead to addiction (Leouzon et al., 2019). Still, online video games are designed to become addictive (Ko, 2014; Montag et al., 2019a; Shi et al., 2019; Sussman et al., 2018) and research has identified negative consequences (e.g. lower quality of life, sleep deprivation, anxiety, depression) related to excessive video gaming (Kuss et al., 2021; Kuss & Griffiths, 2012; Musetti et al., 2019; Sussman et al., 2018).

### Prevalence and Correlates of Internet Addiction and Gaming Disorder

A recent systematic review and meta-analysis examined 133 epidemiological studies (from 31 nations) reporting prevalence rates of what the authors have called *generalized internet addiction* (GIA) or *internet gaming disorder* (IGD) (Pan et al., 2020). The weighed average prevalence for GIA was 7.02% (95% CI, 6.09% - 8.08). Published prevalence rates for specific subpopulations tend to be higher. For example, a meta-analysis of studies comprising a total of 3651 medical students found a pooled prevalence of 30.1% (Zhang et al., 2018). Another meta-analysis of internet addiction among 1,818 healthcare professionals established the pooled prevalence rate at 9.7%. Higher IA scores were associated with emotional or behavior problems, but depressive symptoms were the most common across studies (Buneviciene & Bunevicius, 2021).

When compared to unspecified internet addiction, the prevalence of IGD tends to be lower. It was estimated at 2.47% (95% CI, 1.46% - 4.16%) in the Pan et al. (2020) meta-analysis, 4.6% (95% CI, 3.4% - 6.0%) in the adolescent population (Fam, 2018), and 10.1% (95% CI, 7.3% - 13.8%) in Southeast Asia (Chia et al., 2020). Contrary to the popular image of children and adolescent spending an excessive amount of time playing video games, the latter meta-analysis found that “samples involving older participants showed higher prevalence rate than those involving younger individuals.” (Chia et al., 2020, p.1). Prevalence of IGD was also higher for males (Fam et al., 2018).

### Autism Spectrum Disorder and Problematic Internet Use

Although anyone with access to ICT could develop internet addiction or gaming disorder, core features of autism spectrum disorder (ASD) may place this population at an increased risk. ASD is a neurodevelopmental disorder characterized by two main categories of symptoms: 1- persistent deficits in social communication and social interaction across multiple contexts; 2- restricted, repetitive patterns of behavior, interests, or activities. Impairments in social communication and interactions might make online interactions more comfortable than face-to-face face (Benford & Standen. 2009; Gillespie-Lynch et al., 2014; Roth & Gillis, 2015; Sallafranque Saint-Louis & Normand, 2017), leading autistic individuals to favour online chatting, texting, and gaming to in-person conversation or play. Research has also pointed to an exceptional affinity of individuals with ASD to use technology (Kuo et al., 2014; Mazurek et al., 2012; Orsmond & Kuo, 2011; Shane & Albert, 2008; Wei et al., 2013) or favour access to an electronic device as a reinforce over an edible item (Scheithauer et al., 2022). Therefore, online-related activities may fit their restricted interests, and their repetitive behaviors, impulsivity and difficulties with response inhibition (Corbett et al., 2009) could make it difficult to quit. Considering these characteristics, some researchers believe that individuals with ASD are at increased risk of developing an internet addiction or gaming disorder (Lane & Radesky, 2019; MacMullin et al., 2016; Roth & Gillis, 2015).

Another risk factor for individuals with ASD to become addicted to video games or the internet could be the comorbidities of ASD with attention deficit disorder, anxiety or depression (Lai et al., 2019). Each of these mental health conditions has been associated with internet addiction (Brooks & Longstreet, 2015; Czinz & Hechanova, 2009; De Vries et al., 2018; Romano et al., 2013; 2014; Seki et al., 2019; Sussman et al., 2018) or excessive gaming (Arcelus et al., 2017; Gentile et al., 2011; Ko et al., 2012; Sussman et al., 2018).

#### Internet Addiction

While some studies have shown a positive correlation between autistic traits in young adults and internet addiction (Arcelus et al., 2017; De Vries et al., 2018; Finkenauer et al., 2012; Romano et al., 2014; Shane-Simpson et al., 2016 [Study 1]), few have actually measured internet addiction in clinical samples of individuals diagnosed with ASD. Indeed, a recent systematic review of problematic internet use among the autistic population (Murray et al., 2021) identified four child or adolescent studies (Coskun et al., 2020; Kawabe et al., 2019; So et al., 2017; 2019) and only one adult study (Shane-Simpson et al., 2016 [Study 2]) with estimated prevalence rates of IA based on clinical samples of autistic individuals. Using Young’s *Internet Addiction Test* (Young, 1998), the estimated prevalence of “possible” internet addiction in autistic children and adolescents ranged between 38.3% and 49.4%. When the more severe cut-off for IA is applied, the prevalence rates drop to 10.8 % (So et al., 2017), 8.9% (So et al., 2019) and 5% (Coskun et al., 2020), which is similar to the prevalence of IA (6.09% - 8.08%) in the general population (Pan et al., 2020).

The adult study used a different measure of PIU, the *Compulsive Internet Use Scale* (Meerkerk et al., 2009). Based on a sample of US college students with a declared diagnosis of ASD (*N* = 33) compared to a matched control group (*N* = 33), an identical proportion of 8/33 (24%) met the criteria for “high compulsive internet use” among autistic and non-autistic students (Shane-Simpson et al., 2016). Furthermore, Shane-Simpson and collaborators (2016) tested the relation between problematic internet use and a measure of autistic traits (the Social Responsiveness Scale – A; Constantino & Gruber, 2012), broken down into a social symptoms subscale score and another subscale of restricted interests and repetitive behavior. Although no statistically significant correlation between either subscales and PIU could be detected, they found a positive trend between the Restricted Interest and Repetitive Behavior subscale and PIU (*r*_*s*_ = .38, p = .03).

#### Gaming Disorder

Studies focusing exclusively on problematic gaming among autistic children and adolescents have relied on parent reports (MacMullin et al., 2016; Mazurek & Engelhardt, 2013a,b; Mazurek & Wenstrup, 2013; Paulus et al., 2019). This could lead to an underestimation of this issue in children’s lives, as one of the symptoms of IGD is lying about the amount of time spent online. Therefore one needs to be cautious when interpreting their results. Parents were asked to estimate the amount of time their children spent playing video games, as well as rate the intensity (on a 5-point Likert scale ranging from “Never” to “Always”) of other indicators of problematic gaming. Four of the five studies had a comparison group of non-autistic youth, and all have found higher mean scores indicative of problematic gaming in the group with autism. Autistic children also spent more time gaming in two studies (Mazurek & Wenstrup, 2013; Paulus et al., 2019), and when compared to girls with ASD, boys with ASD spent significantly more time playing video games (MacMullin et al., 2016). Furthermore, scores on problematic gaming measures were correlated with inattention (Mazurek & Engelhardt, 2013 a,b) and opposition (Mazurek & Engelhardt, 2013b).

Despite the fact that the video game industry has recently established the mean age of video game players to be 33 years in the US and that 76% of gamers are over 18 years of age (Entertainment Software Association, 2022), to our knowledge there is only one published study of gaming in autistic adults (Engelhardt et al., 2017). This sample comprised 59 adults with ASD (*M* age = 20.42, SD = 2.01; 86% male), and a comparison group of 60 adults without ASD (*M* = 20.54, SD = 1.34; 87% male). The results showed that male gender was correlated with gaming for both groups, but that adults with ASD displayed more symptoms of gaming disorder. Further, the authors reported that the percentage of free time spent gaming was a better predictor of addiction symptoms than the number of hours spent gaming, regardless of ASD-status.

### Current Study

Experts have argued that individuals with ASD may be at increased risk of experiencing problematic internet use (generalized internet addiction or gaming disorder), yet little research has examined this phenomenon in samples of adults diagnosed with ASD. Given the paucity of data available, the objective of this paper is to examine problematic internet use and gaming from a larger study of substance related and addictive disorders in a sample of adolescents and young adults with ASD. Our specific research questions were:

1. How many individuals with ASD in our sample scored within the clinical range of problematic internet use or gaming disorder? We wanted to compare our data with published prevalence rates among the general population as well as the autistic population.
2. What are some of the correlates of problematic internet use or gaming in young adults with ASD? We wanted to test the association reported in other studies between problematic internet use and age, gender, as well as mental health issues such as attention deficit, depression and anxiety.

## Methods

### Procedure

A convenience sample of individuals with ASD between the ages of 16 and 30 years old was recruited through social media posts and organizations providing services to people with ASD in Canada, following approval by the principal investigator’s university board of ethics. The online survey was designed to measure substance and non-substance use as well as known risk and protective factors to substance related and addictive disorders among individuals with ASD. The advertisement for the study was the following:

**LIFESTYLE AND SUBSTANCE USE OF PEOPLE WITH AUTISM**

Online access

**Goal of the research**

This project aims to describe the lifestyle habits of young adults with ASD (16 to 30 years old) and their using habits of substances (alcohol, drugs), Internet and gambling. Working together, researchers, service providers and stakeholders for people with ASD and people with addictions want to develop a collaboration between their services to better support youth with ASD and addictions. This survey is a first step in the research.

You want to advance knowledge about the lifestyle and using habits of people with autism?

You have a diagnosis of autism spectrum disorder (ASD)?

You are between 16 and 30 years old? Click on the link and complete the survey:

https://is.gd/PAADCanada

CONFIDENTIALITY INSURED

10 gift cards to win among all respondents

Data analyses for the current paper include responses related to sociodemographic characteristics, video game use and internet use. The survey could be completed online only (in approximately 30 minutes) in French or English according to the respondent’s preference. Data collection took place between April and August 2019 (i.e. before the pandemic).

### Participants

Sixty-five individuals with ASD gave their informed consent and completed the survey. Inclusion criteria were that participants were between the ages of 16 and 30, could understand and answer favorably the recruitment ad in French or English, fill out the online survey in either French or English, and had received an official diagnosis of ASD, Asperger syndrome, or pervasive developmental disorder by a professional (e.g., psychologist, psychiatrist, pediatrician, doctor) as declared in item 6 of the demographic data section. Older and younger respondents or those who answered that they had self-diagnosed or were diagnosed by someone in a field other than mental health or the medical profession (e.g. teacher, parent) were not retained for the analyses.

The mean age of participants was 23.77 years (*SD* = 4.3). There were 34 female and 29 male respondents (2 non-responses) according to birth sex. Mean age at diagnosis was 14.74 years (*SD* = 8.06). Two thirds of the participants reported at least one co-occurring mental health condition. The most common conditions were anxiety disorder (38.8%, *N* = 26), attention deficit hyperactivity disorder (ADHD; 35.8%, *N* = 24) and depression (29.9%, *N* = 20). A summary of the sociodemographic characteristics of the sample is presented in Table 1.

**Table 1.** Participant Sociodemographic Characteristics

|  | *Total n = 65* | *%* |
| --- | --- | --- |
| Age group |  |  |
| 16-20 years old | 18 | 27.7 |
| 21-25 | 18 | 27.7 |
| 26-30 | 29 | 44.6 |
| Gender |  |  |
| Male | 27 | 41.5 |
| Female | 33 | 50.8 |
| Non-Binary | 2 | 3.1 |
| Transgender | 2 | 3.1 |
| Fluid | 1 | 1.5 |
| Province |  |  |
| Quebec | 40 | 61.5 |
| British Columbia | 9 | 13.8 |
| Other | 12 | 18.5 |
| Language |  |  |
| French | 40 | 61.5 |
| Highest grade achieved |  |  |
| Elementary school | 12 | 18.5 |
| Secondary school | 18 | 27.7 |
| College | 22 | 33.9 |
| University | 11 | 16.8 |
| Employed | 28 | 43.1 |
| Co-occurring condition |  |  |
| Anxiety | 25 | 38.5 |
| ADHD | 23 | 35.4 |
| Depression | 19 | 29.2 |
| Other | 22 | 33.9 |

### Measures

### Problematic Internet Use

Problematic internet use was assessed using the 5-item version of the *Compulsive Internet Use Scale* (CIUS-5; Besser et al., 2017). This instrument has been validated in both French and English (Lopez-Fernandez et al., 2019) and showed good internal reliability (Cronbach’s *a* = 0.79) with our sample. It had also been used with a sample of autistic adults in another study (Shane-Simpson et al., 2016 [Study 2]). Respondents rated the frequency of each item on a 5-point scale [0 (*never*) to 4 (*very often*)]. Items touch upon time spent online (e.g. How often do you find it difficult to stop using the Internet when you are online? How often do others (e.g., partner, children, parents) say you should use the Internet less?) as well as negative consequences of internet use (e.g. How often are you short of sleep because of the Internet? How often do you neglect your daily obligations (work, school, or family life) because you prefer to go on the Internet?)

Combining response choices *seldom* and *sometimes* to equal a score of 1 and *often* and *very often* to equal a score of 2, a total cut-off score of 9 indicates the need for the individual to be evaluated for internet addiction, with a sensitivity of .96 (Besser et al., 2017).

#### Gaming Disorder

Potential problematic video game use was measured using the *Ten-Item Internet Gaming Disorder Test* (IGDT-10; Király et al., 2017). The IGDT-10 is a short screening instrument developed to assess internet gaming disorder as proposed as an emerging condition in the *Diagnostic and Statistical Manual of Mental Disorders*, fifth edition (DSM-5; APA, 2013). The IGDT-10 contains ten statements corresponding to gaming disorder symptoms, with respondents rating each item on a 3-point scale [0 (*never*), to 2 (*often*)]. Items refer to loss of control in the amount of time spent gaming (e.g. Have you ever in the past 12 months felt the need to play more often or played for longer periods to feel that you have played enough? Have you ever in the past 12 months unsuccessfully tried to reduce the time spent on gaming?), negative consequences (e.g. How often have you felt restless, irritable, anxious and/or sad when you were unable to play or played less than usual? Have you played a lot despite negative consequences [for instance losing sleep, not being able to do well in school or work, having arguments with your family or friends, and/or neglecting important duties]?) and fantasizing about gaming.

Five symptoms scoring 1 or above on the IGDT-10 suggests the presence of a clinical disorder related to internet gaming (Király et al., 2017). This questionnaire has good psychometric properties across several languages (Király et al., 2019), and strong internal reliability in our sample (Cronbach’s *a* = 0.88). Although not specifically designed or adapted for use with an autistic population, a “readability” analysis of the items in French by an online application (https://www.scolarius.com/) indicates that the majority of the items can be understood by persons with primary or secondary education.

## Results

### Problematic Internet Use

The mean score on the CIUS-5 was 5.37 (SD = 2.44), with a minimum of 0 (*n* = 1; 1.5%) and a maximum score of 10 (*n* = 4; 6.2%). Following the suggested cut-off of 9 (Besser et al., 2017), six of 65 participants (9.3%) met criteria to be further evaluated for internet addiction. Chi-square analyses found no significant relations between PIU and gender, age, or co-occurring mental health conditions (anxiety, ADHD, or depression).

### Gaming

Figure 1 shows the distribution of scores on the IGDT-10. More than half (*n* = 37; 57%) of the sample had *never* experienced any of the ten gaming disorder symptoms, and another 40% (*n* = 26) are considered unproblematic with 1-4 symptoms. Only two participants, both 17-year-old males, scored above the cut-off, reporting 8 and 9 symptoms respectively. Again, chi-square analyses found no significant relations between problematic gaming and gender, age, or co-occurring mental health conditions.

**Figure 1.**
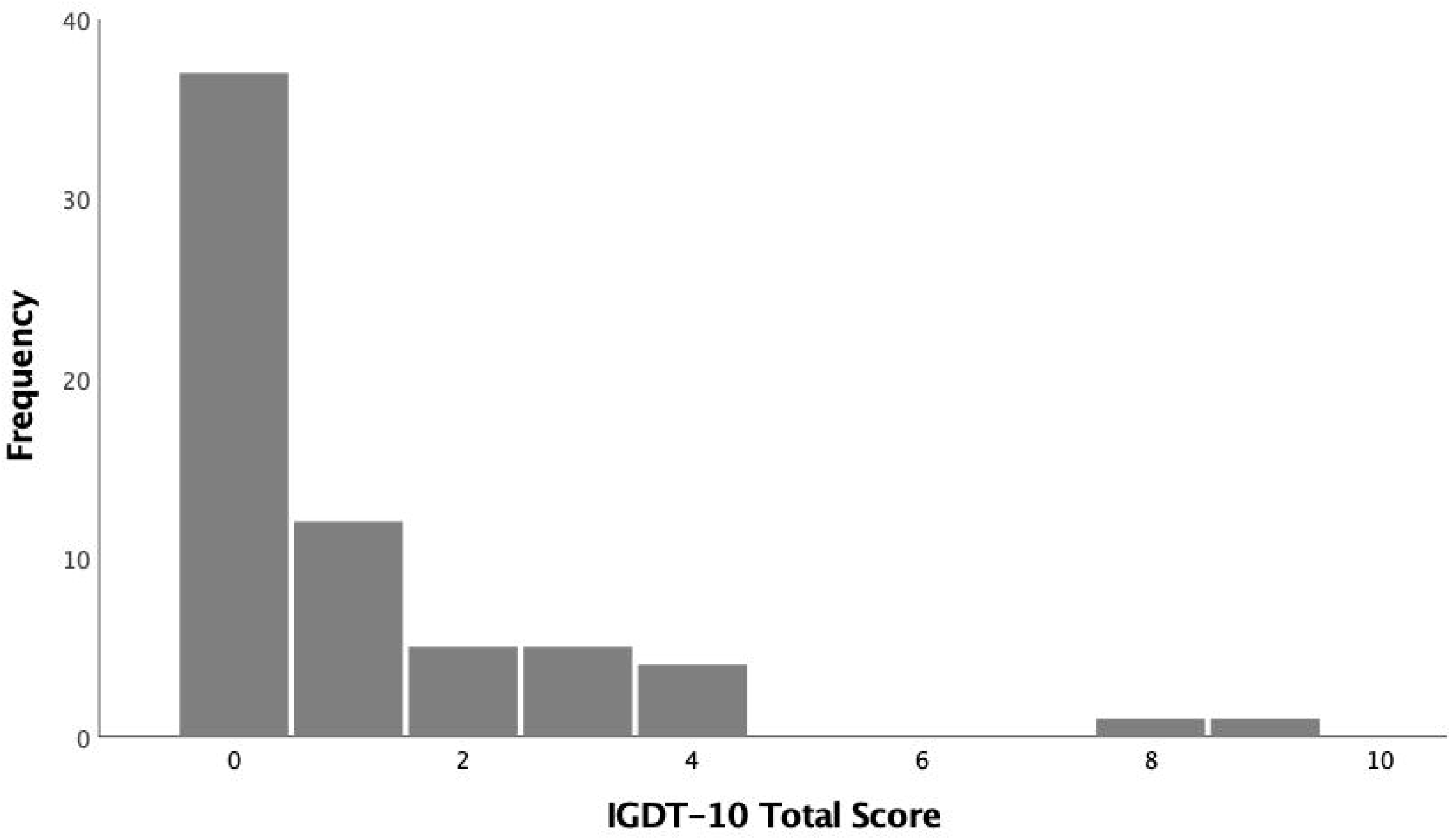
Distribution of IGDT-10 Total Scores

## Discussion

The study explored potential problematic internet use and gaming disorder in a sample of individuals with ASD. Relatively few participants scored high enough to indicate a need to be evaluated for internet addiction or to be given a clinical diagnosis of gaming disorder. Contrary to the hypothesis that individuals with ASD would be more likely to develop an internet or gaming addiction, the proportion of problematic internet users in our sample is very similar to what has been found in the general population (Pan et al., 2020).

First, we identified six (9.3% of the sample) participants with ASD whose symptoms indicated they should be evaluated for internet addiction. This proportion is only slightly higher than the weighed average prevalence of 7.02% found in a meta-analysis of 115 epidemiological studies of IA (Pan et al., 2020), or the proportion of young adults from the general population in Germany (6.7%) identified as problematic internet users (Besser et al., 2017). It is well below the proportion of autistic adults with problematic internet use (24%) reported by Shane-Simpson and collaborators (2016). This divergence might be explained by differences in sample characteristics. Our sample was more heterogeneous in age, level of education and occupation than Shane-Simpson and colleagues’ (2016), who recruited college students with and without ASD enrolled in psychology. Perhaps more noteworthy, they found no significant difference in problematic internet use between groups, indicating once again that autistic adults may be no more at risk of internet addiction than the non-autistic. Unfortunately, because comparable studies (Besser et al., 2017; Shane-Simpson et al., 2016) used the original 21-item version of the CIUS (Meerkerk et al., 2009), sample mean scores cannot be compared with our study results.

As for gaming disorder, only two young adults (3%) out of 65 displayed more than five symptoms on the IGDT-10, suggesting a gaming disorder. These findings are similar to the only other comparable study of problematic gaming in adults with ASD. Although they used a different measure of gaming disorder, Englehardt et al. (2017) found three young adults with ASD (5%) among their 59 participants who met criteria for gaming disorder. Király and colleagues (2019) compared scores on the IGDT-10 in the general population from seven countries/languages and found that the proportion of each sample that met the cut-off score varied between 1.61% and 4.48%, whereas a more recent meta-analysis estimated this prevalence (from 16 studies using various measures of GD) to be 2.47% in the general population (Pan et al., 2020). Together, these results indicate that although individuals with ASD display characteristics that were thought to increase the risk of developing a gaming disorder, the prevalence of gaming disorder in autistic adults may be quite low, and no higher than in the general population. Further, consistent with clinical and scientific evidence indicating that excessive gaming may be a phase that peaks in late adolescence and wanes in adulthood (Leouzon et al., 2019), the problematic gamers in our sample were among the youngest respondents. This goes against findings from a meta-analysis of gaming disorder studies in Southeast Asia, whereby the proportion of GD was higher in samples with older participants (i.e. adults), compared to studies with child and/or adolescent samples (Chia et al., 2020). This could be due to parents exercising greater control over the time that younger video game players spend on their electronic devices, or to the data relying on parent reports, which may underestimate the time their children spend gaming. Future research should explore the presence of gaming disorder among larger samples of individuals with ASD to determine whether age or a particular developmental period is related to gaming disorder.

Finally, more participants indicated potential internet addiction compared to gaming disorder, as was predicted by previous meta-analyses (Chia et al., 2020; Pan et al, 2020). One other study in ASD research has measured and compared internet use and gaming within the same sample. MacMullin et al. (2016) compared parent responses on the 21-item version of the CIUS about their children with ASD to responses from parents of a matched control group without ASD. Parents first answered all items in relation to internet use, then the questionnaire was repeated with the word “internet” replaced with “video game”. Similar to our results, these authors also found a greater proportion of problematic internet use, and a smaller proportion of problematic gaming, in both groups.

## Limitations

This study has limitations that should be addressed in future research. First and foremost are the limitations with regards to the sample of respondents. The current study had a relatively small sample of 65 participants, although this sample size is similar to that of other studies evaluating problem internet use among individuals with ASD (e.g. Coskun et al., 2020; Engelhardt et al., 2017; Kawabe et al., 2019; Shane-Simpson et al., 2016). The fact that participants had to read and understand the recruitment advertisement in order to participate, then answer all questions online, we have certainly not reached autistic youth and adults with greater support needs. Our sample is therefore limited to high or well functioning individuals. Indeed, we lack information about the severity and specificity of autistic symptoms that could be related to the intensity of internet use, or the development of addiction symptoms. We were also unable to confirm the diagnosis of ASD by a trained professional. Hence our results are likely not generalizable to the larger population of adults with ASD.

Given the broader purpose of the survey and recruitment efforts related to identifying individuals with psychoactive substance use and addictive disorders, it is possible that potential participants may have felt the study did not pertain to them. It is also unclear how using the shorter version of the CIUS may have affected the results and we are not able to compare the results to other published studies that used the 21-item version (MacMullin et al., 2016; Shane-Simpson et al., 2016). With only two cases of possible gaming disorder, we lacked the statistical power to analyze any relation between this behavior and other variables identified as correlates in previous studies. Finally, the absence of a matched comparison group of adults without ASD limits our interpretation of group mean and proportion of cases.

## Implications

Clearly more research is needed on larger samples of autistic adults and matched comparison groups before we can refute the theoretical reasons to believe that individuals with ASD are at higher risk of internet addiction and gaming disorder. Measures of autistic traits in the general population indicate that individuals with more autistic-like traits score higher on IA and GD measures. Other studies of clinical samples have also found a higher risk of IA or GD when compared to non-autistic peers (Normand et al., 2021). Given that problematic internet use is associated with symptoms of depression, inattention, impulsivity, hyperactivity and opposition in children and adolescents with ASD (Kawabe et al., 2019; Mazurek & Engelhardt, 2013a,b), these findings need to be explored in an adult sample. Finally, although number of hours online is related to problematic internet use (Engelhardt et al., 2017; Kawabe et al., 2019; MacMullin et al., 2016; Mazurek & Engelhardt, 2013), the percentage of free time adults with ASD spend online is a better predictor of problematic gaming (Engelhardt et al., 2017) and should be taken into account in future research.

## Conclusion

Although ASD is often associated with co-occurring diagnoses of mental health problems and difficulties in coping (Matson, 2016), very little attention has been paid to substance related and addictive disorders (Haasbroek & Morojele, 2021; Ressel et al., 2020) in research and clinical practice. Given the lack of research, clinicians may not be aware of the risks and those in need of support or treatment for addiction have difficulty receiving it. This study, then, contributes to the small body of knowledge currently available in the field and raises questions that should inform further investigation.

## Data Availability

All data produced in the present study are available upon reasonable request to the authors.

